# Post-Ictal Sleep Changes in Human Focal Epilepsy

**DOI:** 10.1101/2025.01.28.25321285

**Authors:** Vaclav Kremen, Yurui Cao, Vaclav Gerla, Filip Mivalt, Vladimir Sladky, Erik St.Louis, Mark Bower, Ben Brinkmann, Kai Miller, Jamie VanGompel, Mark Cook, Tim Denison, Kent Leyde, Gregory A. Worrell

**Affiliations:** Department of Neurology, Mayo Clinic, Rochester, MN, USA; Czech Institute of Informatics, Robotics, and Cybernetics, Czech Technical University in Prague, Prague, Czech Republic; Department of Electrical & Computer Engineering, University of Illinois, Urbana Champaign, IL, USA; Department of Neurology, Yale University, New Haven, CT, USA; Department of Neurologic Surgery, Mayo Clinic, Rochester, MN, USA; Royal Melbourne, Melbourne, Australia; Department of Engineering Sciences, Oxford University, Oxford, UK; Cadence Neuroscience, Seattle, WA

**Author notes:** These authors contributed equally to this work (Vaclav Kremen, Yurui Cao, Vaclav Gerla).

**Keywords:** Focal epilepsy, Slow wave sleep, Local field potentials, Sleep-wake states, Sleep architecture, Seizure-related consolidation

## Abstract

Bidirectional interactions between sleep, seizures, and epilepsy remain incompletely understood. Evidence from animal models and people with focal epilepsy suggest that seizures may engage mechanisms of memory consolidation to reinforce and strengthen synaptic connections within the pathological networks that generates seizures, termed seizure-related consolidation (SRC). Human studies of SRC, however, are limited by small sample size and restricted observations of post-ictal sleep. Using continuous local field potential (LFP) recordings from novel investigational devices implanted in 11 people with drug-resistant focal epilepsy living in their natural environments, we used automated methods to create accurate sleep-wake and seizure catalogs for investigating the interplay between seizures and sleep. Our findings demonstrate that post-ictal slow-wave sleep duration, slow-wave LFP spectral power and waveform slope are increased compared to inter-ictal nights without preceding seizures. The most significant changes localize to the epileptogenic networks generating the subjects’ habitual seizures. These results reveal parallels between SRC and physiological memory consolidation, providing new insights into the role of post-ictal sleep in epilepsy and underscoring the potential of targeting post-ictal sleep for therapeutic interventions in drug-resistant focal epilepsy.

## Introduction

Sleep is essential for maintaining brain health, immune function, and facilitating learning and memory (Huber et al. 2004; Klinzing, Niethard, and Born 2019; Buysse 2014). Chronic sleep disturbances are strongly associated with neurological and psychiatric disorder (Freeman et al. 2020; Ju, Videnovic, and Vaughn 2017). However, determining whether sleep changes directly contribute to brain dysfunction or merely exacerbate disease symptoms remains challenging. Leveraging long-term monitoring in individuals living in their natural environment has recently strengthened the connection between poor sleep and chronic medical conditions, underscoring the potential of sleep as a therapeutic target (Zheng et al. 2024).

In epilepsy, the interplay between seizures and sleep has long intrigued clinicians and researchers (Pavlides and Winson 1989; Gowers 1885). However, most studies utilizing invasive brain recording are constrained to relatively short, multi-day inpatient polysomnography recordings that generate more limited datasets for investigating the interplay between seizures and sleep (Peter-Derex et al. 2020). Despite these limitations, growing evidence supports a reciprocal relationship where seizures disrupt sleep architecture, while poor sleep exacerbates seizure activity (Mark Bower 2024; Malow 2004; Bazil 2017).

Beyond acute seizure events, long-term pathological changes in synaptic connectivity—a hallmark of epileptogenesis—are believed to be a central mechanism in epilepsy progression (Pitkänen et al. 2015). The concept of seizure-related consolidation (SRC) posits that recurrent seizures exploit the mechanisms of memory formation, acting as powerful “engrams” to strengthen pathological networks (Goddard and Douglas 1975; Mark Bower et al. 2017). Evidence of memory reactivation and consolidation has been observed in individuals with temporal lobe epilepsy, further supporting parallels between SRC and physiological memory mechanisms (Mark Bower et al. 2017, 2015). Electrophysiological biomarkers, including inter-ictal epileptiform spikes (IES), high-frequency oscillations, and seizures, may represent powerful, pathological epileptic engrams. These biomarkers are increasingly seen as potential targets for therapeutic interventions aimed at disrupting SRC processes (Hsu et al. 2008; Mark Bower et al. 2015; Kucewicz et al. 2024).

While promising, the investigation of SRC and its relationship with sleep in ambulatory humans is hindered by the inaccuracy of self-reported seizure and sleep diaries (Lauderdale et al. 2008; Elger and Hoppe 2018). To address these challenges, we developed machine learning (ML) frameworks for classifying sleep-wake states and detecting seizures in continuous local field potential (LFP) recordings. The ML algorithms are applied to long-term LFP data from investigational devices implanted in individuals with focal epilepsy and provide accurate sleep-wake and seizure classification catalogs (V. Kremen et al. 2019; Sladky et al. 2022; Mivalt et al. 2022). We previously used the ML algorithms to demonstrate the role of sleep-wake states in multiscale cycles of seizures, inter-ictal epileptiform spikes, mood, impedance, and seizure forecasting (Gregg et al. 2020; Dell et al. 2021; Mivalt, Kremen, et al. 2023; Balzekas et al. 2024).

A particularly interesting finding in our previous studies was that individuals experience longer sleep durations on nights following seizures (Dell et al. 2021). Given sleep’s established role in learning and memory (Diekelmann and Born 2010; Walker and Stickgold 2010), this aligns with the hypothesis that post-ictal sleep facilitates the SRC process. Scalp EEG studies are also consistent with this observation, demonstrating increased post-ictal non-REM (NREM) sleep duration, slow-wave activity (SWA), and waveform slope following focal to bilateral tonic-clonic seizures (Boly et al. 2017).

In the current study, long-term, intracranial LFP recordings from people with focal epilepsy implanted with novel investigational devices show increased post-ictal slow-wave sleep duration, SWA spectral power and waveform slope. These results highlight the potential of targeting post-ictal NREM sleep to disrupt SRC in drug-resistant focal epilepsy.

## Methods and materials

### Human Subjects

We analyzed two cohorts (cohorts #1 & #2) of human subjects with drug-resistant focal epilepsy implanted with investigational devices providing continuous local field potential (LFP) sensing from multiple brain regions.

Cohort #1 (NC) includes continuous, long-term LFP data from subjects implanted with the investigational seizure advisory system (NeuroVista Inc.) between March 24, 2010, and June 21, 2011 in Melbourne, Australia (Cook et al. 2013). Seizure onset networks included neocortical temporal, frontal, and parietal brain regions (supplementary Table 1). Subjects were implanted with four subdural, 4-contact, cortical strip electrode arrays over the region of the seizure focus. The subdural arrays were connected to lead extensions tunneled down the neck to an implantable subclavicular LFP recording and telemetry unit. The LFP data were wirelessly transmitted to a hand-held device running algorithms to detect and forecast seizures (Cook et al. 2013). A subset of six patients from the NC cohort were identified with at least 20 days of continuous inter-ictal and post-ictal LFP data with over 80% of the LFP data captured in both inter-ictal (seizure-free for 48-hours) and post-ictal (seizures preceding) days and nights.

Cohort #2 (mTLE) includes continuous, LFP data from 5 subjects in a clinical trial using the investigational Medtronic Summit RC+S^TM^ between July 10, 2019 and October 23, 2023 at Mayo Clinic, MN, USA (Vaclav Kremen et al. 2024). All subjects had bilateral amygdala-hippocampus onset seizures (supplementary Table 1). Four leads with 4-electrode contacts were targeted at bilateral amygdala-hippocampus and anterior nucleus of thalamus. Lead extensions were tunneled down the neck to a subclavicular rechargeable, electrical stimulation, LFP sensing, and wireless telemetry device.

The neocortical epilepsy subjects (NC 1-6) had electrode contacts in the seizure onset zone (SOZ) and neocortex outside the seizure onset zone (NSOZ). The subjects with mTLE (mT 1-5) had bilateral independent seizures recorded from electrodes in epileptogenic hippocampus bilaterally. Data from 11 subjects was used for the evaluation of the sleep duration, architecture, and slow-wave LFP spectral power and waveform slope. We analyzed SOZ and NSOZ contacts to investigate electrophysiological difference in the brain regions generating seizures in the neocortical epilepsy subjects.

### Intracranial Electrophysiology Recordings

The LFP were continuously recorded from multiple brain structures using the NeuroVista and RC+S^TM^ devices (Figure 1). The NeuroVista device recorded 16 channels of subdural LFP data using an average reference (calculated from the 16 contacts) and sampled at 400 Hz (bandwidth 0.85 – 100 Hz) from temporal, frontal, and parietal neocortex.

**Figure 1:**
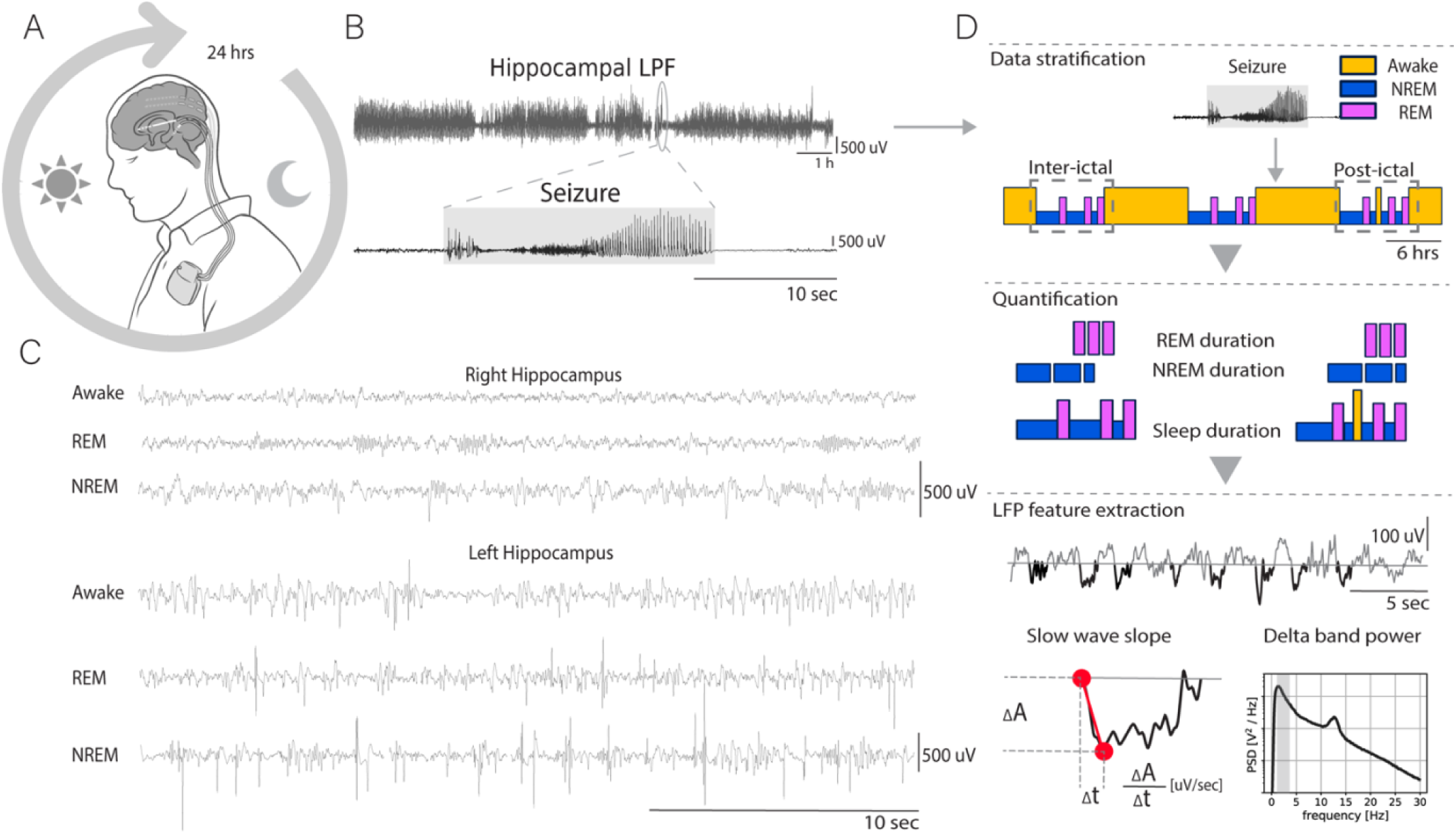
Continuous long-term monitoring of local field potentials (LFP) in people with drug resistant focal epilepsy. A) Investigational implantable neural sensing devices were used to record continuous long-term LFP from amygdala-hippocampus in 5 subjects with mesial temporal lobe epilepsy (mTLE) and in 6 subjects with frontal, temporal or parietal neocortical epilepsy. B) Validated automated machine learning algorithms were used to detect electrographic seizures and to classify each 10 second window of continuous LFP as inter-ictal (non-seizure) or ictal (seizure). C) Sleep-wake state was classified as wakefulness (W), rapid eye movement (REM) or non-REM (NREM 1,2,3) sleep. D) The data were split into nights after seizures (post-ictal) and nights without prior seizures in last 48-hours (inter-ictal). The duration of wakefulness, NREM, and REM and slow-wave sleep activity LFP micro-sleep features were analyzed.

The RC+S^TM^ device recorded four bipolar LFP channels sampled from the 16 parenchyma electrode contacts in the bilateral anterior nucleus of the thalamus (ANT), amygdala (AMG), and hippocampus (HPC). The LFP was sampled at 250 Hz (bandwidth 0.85 – 80 Hz). Long-term recordings from the 5 subjects included at least 1 month of LFP data.

### Automated Seizure Detection

We used an automated deep learning model (convolutional neural network with long short-term memory: CNN-LSTM) to label candidate seizures (Sladky et al. 2022). The CNN-LSTM seizure detector was developed and validated previously within a complex study with multiple sensing devices implanted in naturally occurring canine and human epilepsy. Long-term records from humans and canines with epilepsy living in their natural environments provided an unprecedented data set of spontaneously occurring seizures. These datasets are highly imbalanced because seizures are relatively rare events when compared to the continuous inter-ictal LFP data. The CNN-LSTM seizure detector represents an analysis evolution from early feature-based seizure detectors (Brinkmann et al. 2016) to ML seizure detectors (Nejedly et al. 2019; Sladky et al. 2022). In the current study, all candidate seizure detections were visually reviewed by an epileptologist to create accurate training, validation and testing datasets.

### Automated Behavioral State Classification

To perform sleep-wake classifications over long-term datasets, we used our previously developed ML framework for LFP-based, automated behavioral state classification (V. Kremen et al. 2019, 2017; Mivalt et al. 2022). We have previously validated the sleep-wake classifications against gold standard scalp polysomnography. Our ML framework enables unsupervised, semi-supervised, and supervised learning to classify behavioral states including AWAKE, NREM 2&3, and REM states. The general framework uses human expert-in-the-loop machine learning with training and testing on invasive LFP recordings combined with simultaneous polysomnography (V. Kremen et al. 2019) to AASM criteria (Silber 2012). The same sleep scoring results and the classifier was used in prior NC cohort (Dell et al. 2021; Payne et al. 2021) and RC+S^TM^ (Mivalt et al. 2022; Mivalt, Sladky, et al. 2023) analysis.

### Statistics

Differences between data groups measured in several conditions: SOZ versus NSOZ networks, nights following the seizure (post-ictal nights) versus nights without preceding seizures (inter-ictal nights) were tested using Kruskal-Wallis ANOVA given it is a non-parametric statistical test that avoids assumptions of distribution normality in the measures. Statistical significance for all tests was defined by p-value *<* 0.05. When comparing grouped data, because of the significant variability in values between subjects, we normalized the data from each subject by z-score (*Z* = (*x* −*µ*)/*σ*), where µ is the mean and *σ* is the standard deviation, to avoid bias caused by large absolute values of individual subjects.

### Post-ictal and Inter-ictal Sleep Analysis

To study the alterations in sleep dynamics in NC cohort related to the subjects’ habitual seizures, we required subjects to fulfill the following conditions: **1)** The subject had at least 20 nights of inter-ictal data (defined as nights occurring at least ±48 hours after preceding seizure events). **2)** The subject had at least 20 nights of post-ictal data (defined as nights following seizures occurring during wakefulness). **3)** Each 24-hour data segment contained more than 80% of available data. These data were used to analyze sleep architecture and electrophysiological properties of inter-ictal and post-ictal sleep. Ultimately, the criteria above yielded datasets from 6 subjects out of the original 15 subjects implanted with the NeuroVista device. Inter-ictal nights (no seizures during the preceding 48 hours) and post-ictal nights (seizures during the preceding day) were balanced, with 20 nights from each category for each patient. In the NC cohort, we separated electrodes into two groups: Seizure Onset Zone (SOZ) electrodes versus non-SOZ (NSOZ) electrodes. In total, we analyzed 240 day-nights of chronic LFP recording.

## Results

We analyzed two unique long-term LFP datasets from 11 subjects with focal drug-resistant mesial temporal lobe epilepsy (mTLE) and neocortical epilepsy (NC) (supplementary Table 1)

**Table 1:** Subject Demographics. Abbreviations: ASM = Anti-seizure medications, CBZ=carbamazepine, CLZ=clobazam, CZP=clonazepam, LCM = lacosamide, LEV=levetiracetam, LTG=lamotrigine, OXC=oxcarbazepine, PHT = phenytoin, RTG=retigabine, ZNS=zonisamide, SOZ = Seizure onset zone Seizure onset zones: mTL – mesial temporal lobe, nFT – neocortical frontotemporal, nOP – neocortical occipitoparietal, nPT – neocortical parietal-temporal, nTL – neocortical temporal lobe.

| Subject | Age (yrs) | Sex | Onset (yrs) | ASM | SOZ | #Days/#Seizures |
| --- | --- | --- | --- | --- | --- | --- |
| **Neocortical Epilepsy (Seizure Advisory System, NeuroVista Inc.)** |  |  |  |  |  |  |
| **NC-1** | 35-40 | M | 5-10 | CBZ, LCM, PRP, TPM | nTL; prior ATLE | 465/161 |
| **NC-2** | 45-55 | M | 15-20 | CBZ, CLZ, LEV, LCM | nTL, prior ATLE | 534/1839 |
| **NC-3** | 40-45 | M | 20-25 | LTG, LCM, PHT, RTG | nTL | 709/1845 |
| **NC-4** | 50-55 | F | 15-20 | LEV, OXC, ZNS | FT | 318/378 |
| **NC-5** | 45-50 | M | 20-25 | CBZ, LEV | FT | 355/1425 |
| **NC-6** | 40-45 | M | 10-15 | LCM, LTG, OXC, VPA | OP | 607/264 |
| **Mesial Temporal Lobe Epilepsy (RC+S™, Medtronic Plc.)** |  |  |  |  |  |  |
| **mT-1** | 55-60 | F | 5-10. | GBP, TZP, LGT | mTL | 831/542 |
| **mT-2** | 20-25 | F | 5-10 | LCM, CZP, | mTL; prior ATLE | 392/15 |
| **mT-3** | 40-45 | F | 30-35 | CZP, LEV, CNB | mTL | 401/59 |
| **mT-4** | 35-40 | F | 0-5 | OXC, LEV | mTL | 300/250 |
| **mT-5** | 30-35 | M | 20-25 | VPA, LCM | mTL | 325/219 |

### Seizures in Ambulatory Subjects with Focal Epilepsy

The 11 subjects had seizures originating from hippocampus (5/11), temporal neocortex (3/11), frontal neocortex (2/11), or parietal neocortex (1/11) (supplementary Table 1.). The circadian-ultradian distribution of seizures varied across subjects. Seizures were classified into diurnal (daytime: 8AM – 12-midnight), nocturnal (>12midnight – 8AM), or mixed. The patients with mTLE had mostly diurnal seizures with subject specific unimodal, bimodal or uniform distribution patterns. The patients with neocortical epilepsy (Karoly et al. 2021, 2016) have a more varied temporal distribution of seizures occurrence with both nocturnal and diurnal (Figure 2. A&C).

**Figure 2:**
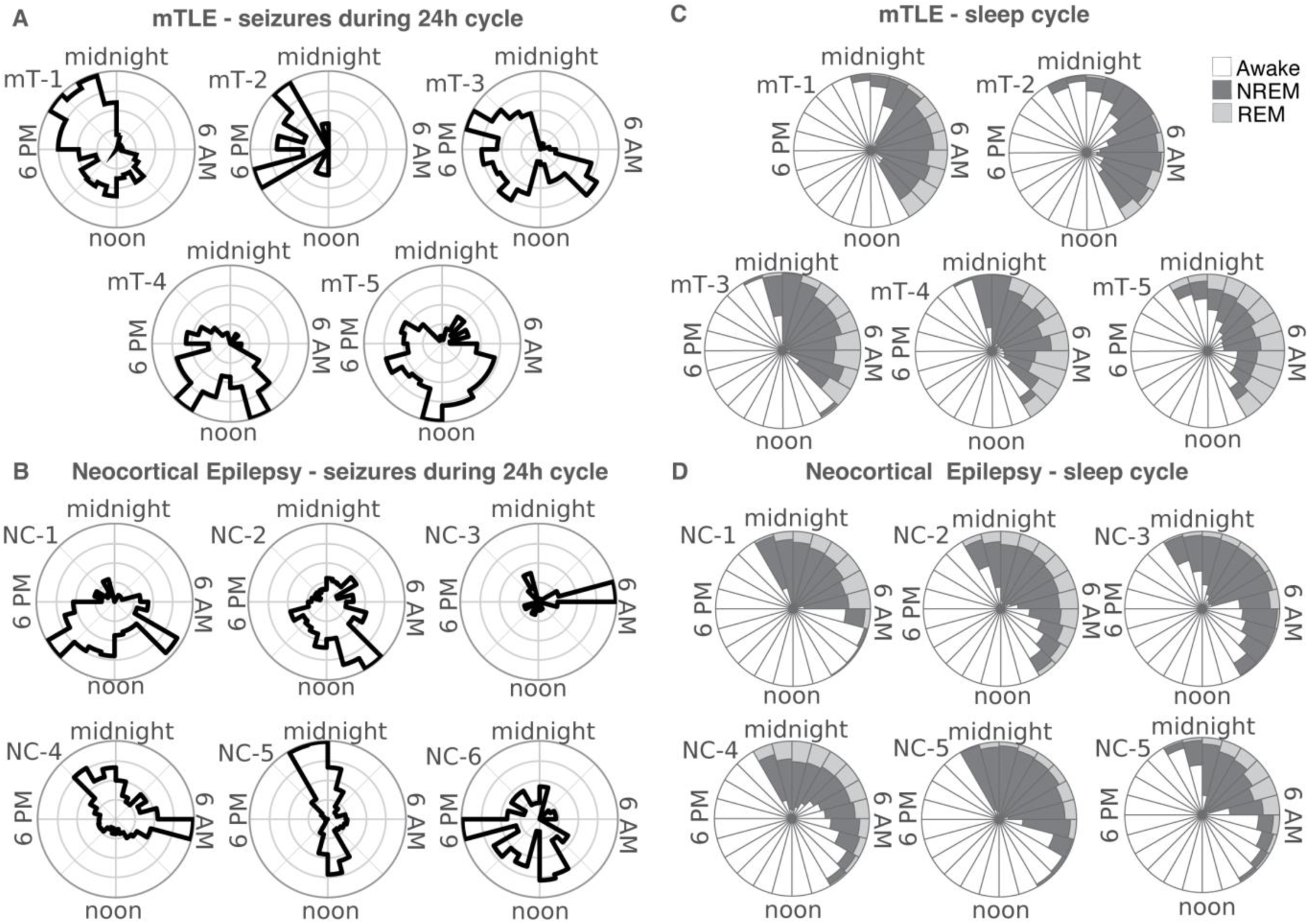
Circadian distribution of seizures and sleep-wake using continuous long-term local field potentials (LFP) recorded from subjects with mesial temporal epilepsy (mTLE) or neocortical focal epilepsy (NC) over months. **A)** 5 subjects with mTLE (mT 1-5) and seizure recorded from amygdala-hippocampus. Seizures are diurnal with seizure occurring at patient specific time-of-day. **B)** Subjects with focal neocortical seizures recorded from temporal (NC 1-3), frontal (NC 4-5) and parietal (NC-6). Two subjects had nocturnal seizures (NC-3&5) with decreased REM during the night **C&D)** Circular histograms of sleep-wake states for mTLE and NC patients (frontal, temporal and parietal).

### Sleep-wake Classification in Ambulatory Subjects with Focal Epilepsy

The circular histograms of the diurnal distribution of behavioral states (Wakefulness, NREM1, NREM2, NREM3, REM) and seizures reveal patient-specific seizure and sleep patterns (Figure 2). Except for subjects mT-2 & NC-3 who had very little REM, the circular histograms for sleep-wake show increased NREM3 in the early part of the night followed by shift to more NREM2/REM cycles with longer REM later in the night, as was reported in limited duration sleep-wake studies (Nir et al. 2011; Ferrara et al. 2012; Hoel et al. 2016; Tononi and Cirelli 2020, 2003). Three subjects with mTLE had markedly reduced REM/NREM ratios (mT-2, NC-3, NC-5). The remaining subjects had more physiologically normal sleep distributions, with more NREM sleep early in the night and more REM later in the night sleep cycle.

### Seizure-associated alterations in sleep architecture and electrophysiology

Analysis of sleep duration (Figure 3) shows the alternation of total sleep duration and NREM sleep in post-ictal versus inter-ictal sleep. The average length of sleep across subjects on inter-ictal nights was 8.3 ± 1.5 hours compared to a longer total sleep time of 8.8 ± 1.4 hours (*p <* 0.05) on nights after seizures occurring during the day and longer post-ictal NREM sleep (5.8 ± 1.2 hours) compared to inter-ictal nights (5.6h ± 1.4 hours; p <0.05).

**Figure 3:**
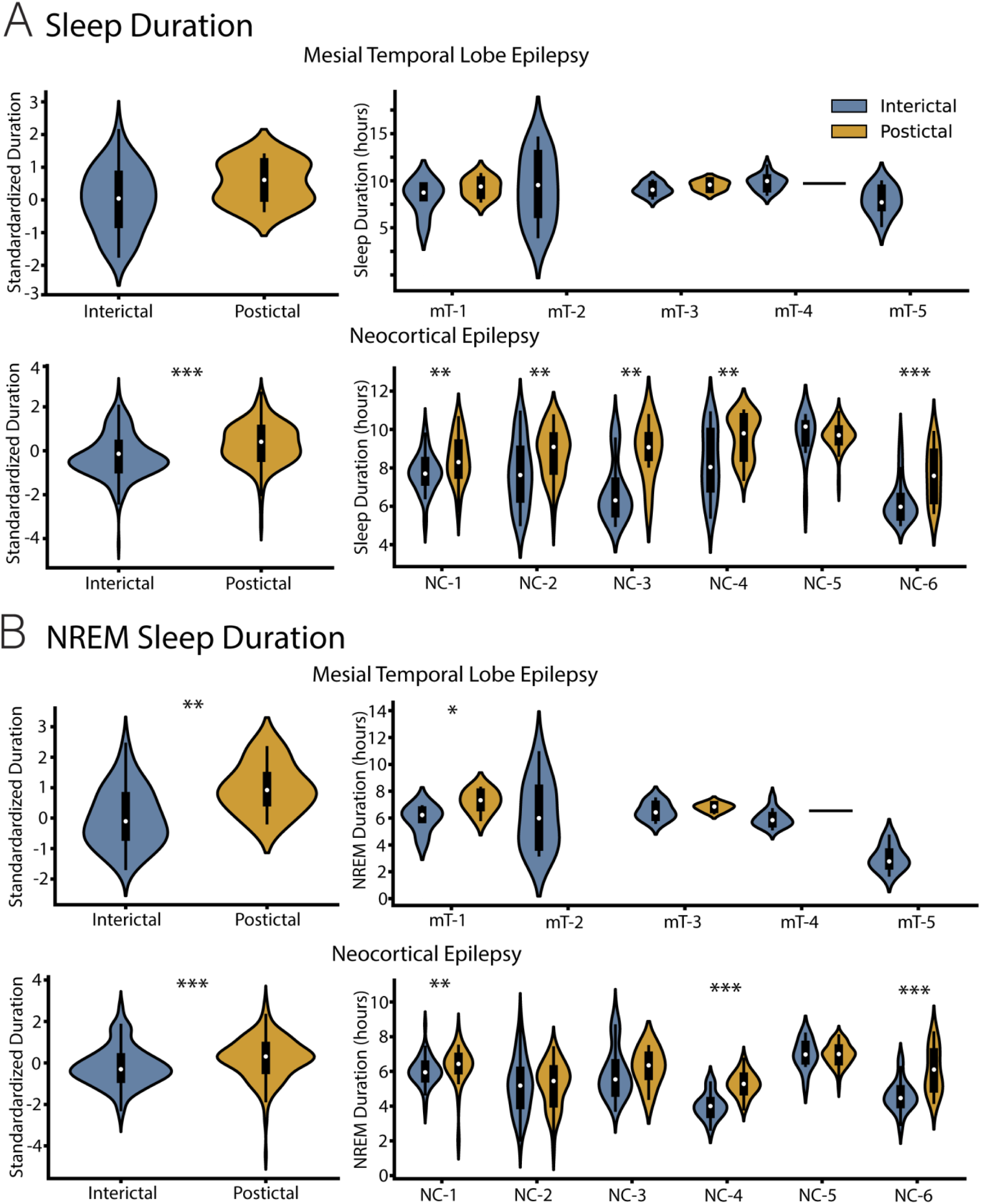
Macro-architecture of inter-ictal and post-ictal sleep. Total sleep duration is the sum of duration of all sleep stages (NREM 1-3 and REM). NREM sleep duration is the sum of all NREM stages. A) Total sleep duration was similar for inter-ictal and post-ictal slow-wave sleep in subjects with mesial temporal lobe epilepsy (mTLE). In subjects with neocortical focal epilepsy (NC) post-ictal sleep was prolonged compared to inter-ictal sleep (p<0.001). On an individual level 83% (5/6 subjects) with NC had increased post-ictal sleep duration. For subjects with mTLE the effects were small with a trend present at a group level. B) The NREM post-ictal sleep was prolonged compared to inter-ictal for both NC and mTLE subjects. On an individual level 50% (3/6 subjects) with NC had increased post-ictal NREM sleep durations. For subjects with mTLE the effects were small and only present at a group level. (* p < 0.05, ** p < 0.01, *** p < 0.001)

There were electrophysiologic changes in post-ictal sleep nights. Both slow-wave sleep activity (SWA: 1 – 3 Hz) power and LFP waveform slope during NREM sleep are increased on post-ictal nights compared to inter-ictal nights. The SWA power is higher in post-ictal sleep for 82% (9/11) subjects overall, 4/5 subjects with mTLE and 5/6 subjects with NC. Furthermore, the increased LFP slow-wave power and slope are greater in the SOZ compared to NSOZ in the neocortical subjects where the analysis is possible (supplementary Table 2). The increased SWA power and slope is bigger during the early night (first four hours of the night) of sleep but persists into the late half of the night. Notably, larger post-ictal effects on SWA power and slope early in the sleep cycle compared to later is consistent with observations from shorter data sets by Boly et al. (Boly et al. 2017).

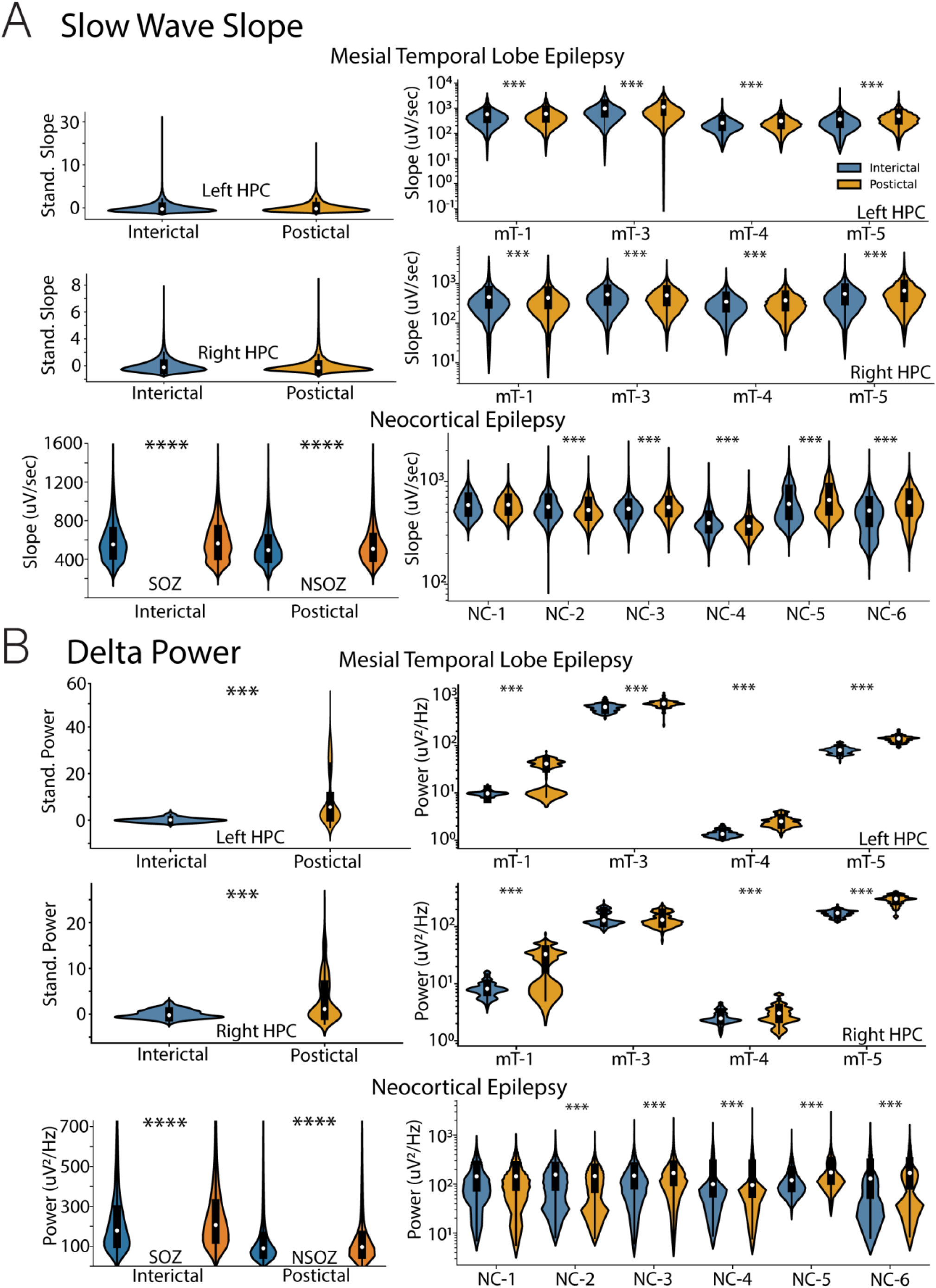
Micro-architecture of sleep in post-ictal nights. Analysis of Local field potentials (LFP) in slow-wave (NREM) sleep in inter-ictal nights and post-ictal nights after seizures. Automated classification of inter-ictal and post-ictal wakefulness and sleep was performed retrospectively using all LFP channels. Inter-ictal night was defined by absence of seizures during the preceding 48 hours. Post-ictal night is defined as sleep after seizures during awake time preceding the night. The LFP slow-wave analysis during NREM sleep was performed to quantify the slow-wave slope **(A)** and delta frequency (1 - 3 Hz) band power **(B)** in each 30 seconds data segment. The LFP slow-wave activity from the amygdala-hippocampal recordings in mesial temporal lobe epilepsy (mTLE) and neocortical subdural LFP recordings in neocortical focal epilepsy (NC) were analyzed independently. **A)** The LFP slow-wave slope was increased during post-ictal nights compared to inter-ictal nights for both mTLE and NC subjects. On an individual level 100% (4/4 subjects) with mTLE and 83% (5/6 subjects) with NC had increased post-ictal slow-wave LFP slopes. **B)** The LFP delta power was increased during post-ictal nights compared to inter-ictal nights for mTLE and NC subjects. On an individual level 100% (4/4 subjects) with mTLE and 83% (5/6 subjects) with NC had increased post-ictal power in delta frequency band. For grouped NC panel, the absolute Delta band power is shown to highlight the differences between the absolute values of power at seizure onset zone (SOZ) and non-seizure onset zone (NSOZ) electrodes. (p < 0.01, *** p < 0.001).

## Discussion

This study investigates post-ictal slow wave sleep using long-term continuous LFP recordings in ambulatory subjects with focal epilepsy living in their natural home environment. We quantified inter-ictal and post-ictal slow-wave sleep and observed consistent changes in post-ictal slow wave sleep (NREM) duration, SWA spectral power, and SWA slope. The results aligned with systems-related consolidation where increases in NREM slow-wave sleep activity are electrophysiological biomarkers of the consolidation process (Klinzing, Niethard, and Born 2019). Similar findings were also reported in scalp EEG studies where NREM SWA power and waveform slope was positively correlated with the frequency of secondary generalized seizures in the days preceding sleep and at scalp locations associated with their focal epilepsy (Boly et al. 2017; Moffet et al. 2020). Focal increases in sleep NREM SWA in the nights after motor learning tasks have also been described in normal subjects (Huber et al. 2004). In summary, the findings presented here support a process of seizure-related consolidation in human focal epilepsy.

Furthermore, recent results from animal models support targeting consolidation to disrupt fear memory consolidation (Clawson et al. 2021) and epileptic networks (Lai et al. 2024). Lai et al. initially showed that an mTLE pathway inhibitor delivered during the post-ictal period yielded the largest seizure and IES reduction, and subsequently demonstrated similar findings for IES in a pilot human trial (Lai et al. 2024). These results support the potential importance of temporal post-ictal targeting of therapy to prevent pathologic learning of epilepsy engrams.

Studies in larger numbers of subjects are required to confirm the preliminary findings reported here. Future studies should also address the relations between sleep patterns and common comorbidities, such as mood and memory dysfunction. Lastly, these findings may prove useful for investigating neuromodulation approaches that attempt to disrupt the consolidation of seizure engrams by targeting post-ictal slow wave sleep.

## Data Availability

All data produced in the present study are available upon reasonable request to the authors.

## Acknowledgments

The authors thank the people with epilepsy for participating in this research. We thank Certicon a.s. for the use of CyberPSG tool for the visual review of EEG.

