## Supplementary Data for "Post-Ictal Sleep Changes in Human Focal Epilepsy"

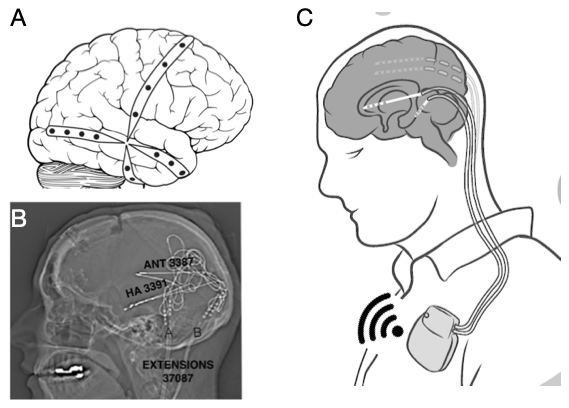

**Supplementary Figure 1:** **Investigational Sensing Devices for Streaming Local Field Potentials (LFP) in focal epilepsy.** **A)** Schematic of NeuroVista Inc. seizure advisory system implant. There are four 4-contact subdural electrode arrays placed in subdural space to record LFP from neocortex. Representative example of subject with temporal lobe epilepsy. **B)** Lateral X-ray of Medtronic Plc. RC+S^TM^ electrode implant in subjects with mesial temporal lobe epilepsy. Each subject had 4-contact penetrating electrode arrays implanted in bilateral amygdala-hippocampus (AH) and anterior nucleus of thalamus (ANT). The figure shows the lead extensions that are tunneled down the neck to a subclavicular pocket. **C)** Schematic of brain LFP wireless streaming devices.

| Subject | Age (yrs) | Sex | Onset (yrs) | ASM | SOZ | #Days/#Seizures |
| --- | --- | --- | --- | --- | --- | --- |
| Seizure Advisory System (NeuroVista Inc.) | | | | | | |
| NC-1 | 35-40 | M | 5-10 | CBZ, LCM, PRP, TPM | nTL; prior ATLE | 465/161 |
| NC-2 | 45-55 | M | 15-20 | CBZ, CLZ, LEV, LCM | nTL, prior ATLE | 534/1839 |
| NC-3 | 40-45 | M | 20-25 | LTG, LCM, PHT, RTG | nTL | 709/1845 |
| NC-4 | 50-55 | F | 15-20 | LEV, OXC, ZNS | FT | 318/378 |
| NC-5 | 45-50 | M | 20-25 | CBZ, LEV | FT | 355/1425 |
| NC-6 | 40-45 | M | 10-15 | LCM, LTG, OXC, VPA | OP | 607/264 |
| RC+S^TM^ (Medtronic Plc.) | | | | | | |
| mT-1 | 55-60 | F | 5-10. | GBP, TZP,LGT | mTL | 831/542 |
| mT -2 | 20-25 | F | 5-10 | LCM, CZP, | mTL; prior ATLE | 392/15 |
| mT-3 | 40-45 | F | 30-35 | CZP, LEV, CNB | mTL | 401/59 |
| mT -4 | 35-40 | F | 0-5 | OXC, LEV | mTL | 300/250 |
| mT -5 | 30-35 | M | 20-25 | VPA, LCM | mTL | 325/219 |

**Supplementary Table 1: Subject Demographics.** Abbreviations: ASM = Anti-seizure medications, CBZ=carbamazepine, CLZ=clobazam, CZP=clonazepam, LCM = lacosamide, LEV=levetiracetam, LTG=lamotrigine, OXC=oxcarbazepine, PHT = phenytoin, RTG=retigabine,ZNS=zonisamide, SOZ = Seizure onset zone Seizure onset zones: mTL – mesial temporal lobe, nFT – neocortical frontotemporal, nOP – neocortical occipitoparietal, nPT – neocortical parietal-temporal, nTL – neocortical temporal lobe.

**Early Night**	**Late Night**

|  | SOZ | | | NSOZ | | | SOZ | | | NSOZ | |
| --- | --- | --- | --- | --- | --- | --- | --- | --- | --- | --- | --- |
| Biomarker | Interictal | Post-Ictal | Interictal | | Post-Ictal | Interictal | | Post-Ictal | Interictal | | Post-Ictal |
| Delta power | 223 ± 165 | 247 ± 168 | 138 ± 147 | | 141 ± 146 | 207 ± 176 | | 233 ± 184 | 103 ± 96 | | 111± 109 |
| Delta slope | 586 ± 208 | 593 ± 214 | 531 ± 190 | | 547 ± 197 | 582 ± 214 | | 573 ± 214 | 525 ± 194 | | 519±183 |

**Supplementary Table 2:** **Changes in Delta band power and slope** in Seizure Onset Zone (SOZ) and Non-Seizure Onset Zone (NSOZ) electrodes and at interictal nights without any seizure the same day versus nights after the seizure. The average ± standard deviation of power in the Delta band and slope of the signal in the Delta band are shown in the table (all comparisons p < 0.01).
